# A digital measure of eye movements during reading sensitively captures oculomotor and speech dysfunction, early changes, and disease progression in ataxias

**DOI:** 10.1101/2025.01.13.25320487

**Authors:** Brandon Oubre, Faye Yang, Anna C. Luddy, Rohin Manohar, Nancy N. Soja, Christopher D. Stephen, Jeremy D. Schmahmann, Divya Kulkarni, Lawrence White, Siddharth Patel, Anoopum S. Gupta

## Abstract

**Objective:** Sensitive behavioral measures are needed for clinical trials in ataxias and other neurodegenerative diseases. We hypothesized that quantitative analysis of eye movements during a natural multi-component task (passage reading) could produce a measure capable of capturing subclinical signs and disease progression.

**Methods:** Binocular gaze sampled at 1000 Hz was collected from 102 individuals with ataxia (including 36 spinocerebellar ataxias, 12 Friedreich’s ataxia, and 5 multiple system atrophy) and 70 healthy controls. Longitudinal data were available for 26 participants with ataxia in the ongoing natural history study. The Reading Eye Abnormality Digital (READ) score was developed by training a regression model to aggregate saccade and fixation kinematics.

**Results:** Mean displacement of fixations, the number and frequency of saccades, and the proportion of regressive saccades were related to oculomotor dysfunction, speech dysfunction, and overall ataxia severity. The READ score was reliable (ICC=0.96, p<0.001) and correlated with Brief Ataxia Rating Scale total score (r=0.82, p<0.001), oculomotor (r=0.52, p<0.001) and speech (r=0.73, p<0.001) subscores, and patient reports of function. The READ score detected subclinical oculomotor (AUC=0.69, p=0.02) and speech (AUC=0.72, p<0.001) signs and disease progression (d=0.36, p=0.03). The Brief Ataxia Rating Scale was less sensitive to progression (d=0.27, p=0.08).

**Interpretation:** Digital measures of eye movements are a promising approach for sensitively measuring ataxia in clinical trials (including early-stage disease) and may have utility in other neurodegenerative diseases affecting speech or ocular control.

**Thumbnail:** 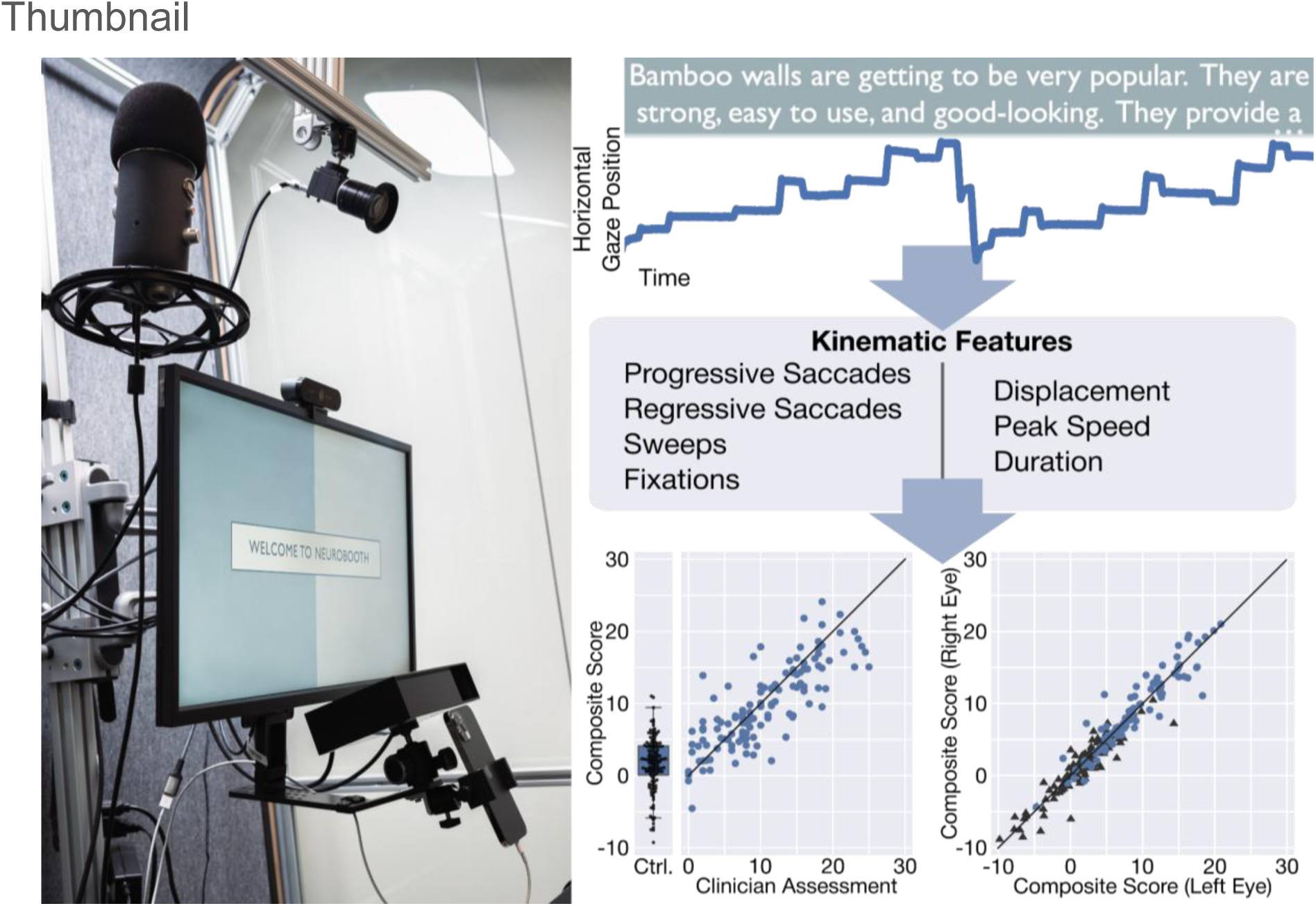

## Introduction

Behavioral measures of disease that capture early changes and progression are needed to support the accelerating drug development efforts in cerebellar ataxias and other neurodegenerative diseases.^1,2^ Semiquantitative clinical rating scales, such as the Brief Ataxia Rating Scale^3^ (BARS), Scale for Assessment and Rating of Ataxia^4^ (SARA), and International Cooperative Ataxia Rating Scale^5^ (ICARS) are currently used in ataxia research and natural history studies.^6–8^ While useful for comprehensively assessing different aspects of the ataxia phenotype, these evaluations were not designed to measure the earliest stages of disease or quantify progression over short time intervals.^7^ Furthermore, these scales isolate each behavioral domain (e.g., speech, limb movement, eye movements), which raises questions about the functional relevance of the individual assessments.^2,8^

Speech and eye movements are key behavioral domains affected in ataxias^9,10^ and other neurodegenerative diseases such as Parkinson’s disease, atypical parkinsonian disorders, Huntington’s disease, and amyotrophic lateral sclerosis.^11,12^ Changes in speech and eye movements occur early in the disease process and progress over time.^9,13–15^ Furthermore, neural control of speech and eye movements are widely distributed across the brain and are influenced by cognitive, affective, and attentional processes in addition to motor processes.^11,12^ The wide distribution of control suggests that changes in eye movements and speech may occur when disease disrupts any level of the system, and may form the basis of highly sensitive disease measures.

Quantitative assessment of speech and eye movements using microphones and eye tracking devices has already shown promise for detecting early disease signs and quantifying progression over time in various neurologic disorders, including ataxias,^14,15^ parkinsonian disorders,^16,17^ and Huntington’s disease.^18,19^ However, the majority of behavioral tasks used for assessing these domains are domain-isolated tasks similar to those included in clinical rating scales. In contrast, recent work by Terao, et al. investigated the interplay between speech and eye movements while reading aloud in a cohort of 16 individuals with cerebellar ataxias and 18 individuals with Parkinson’s disease and found that eye-voice coordination is affected differently by each phenotype.^20^ More specifically, frequent regressions resulted in slower speech for individuals with cerebellar ataxia. This finding, in conjunction with other known changes to eye movement kinematics, supports the use of reading as a cross-domain task for the assessment of ataxias.^20,21^ However, reliability, relationships with clinical severity and patient-report, and progression over time have not been evaluated to understand potential utility as an outcome measure.

In this work, we collected eye tracking data from a large cohort of individuals with ataxia during a reading aloud task to test the hypothesis that eye movement patterns during a multi-component and functionally-relevant task could produce a sensitive and meaningful digital measure of ataxia.

## Materials and methods

### Data Collection

The data used in this analysis were obtained from the Neurobooth^22^ study. Participants typically visited the Neurobooth immediately prior to or after their clinic appointments, at which time neurologist-administered assessments were documented. Participants were seated on either 1) a full-back chair with a standard headrest (Fig. 1A) or 2) a wheelchair. Participants’ heads were located roughly 58 cm (57.8 ± 3.9 cm; mean ± standard deviation) from a 24-inch BenQ monitor. Eye tracking calibration was performed using a five-point stimulus at the beginning of the Neurobooth task battery. For the passage reading task, a static screen displaying the Bamboo passage^23^ (Fig. 1B) was presented after a short instructional video. An EyeLink Portable Duo (SR Research Ltd.) in a head-free configuration recorded binocular gaze data at 1000 Hz during the task performance. Participants took as much time as needed to read the passage aloud (35.6 ± 13.3 s). See the Supplementary Methods for additional protocol details.

**Figure 1.**
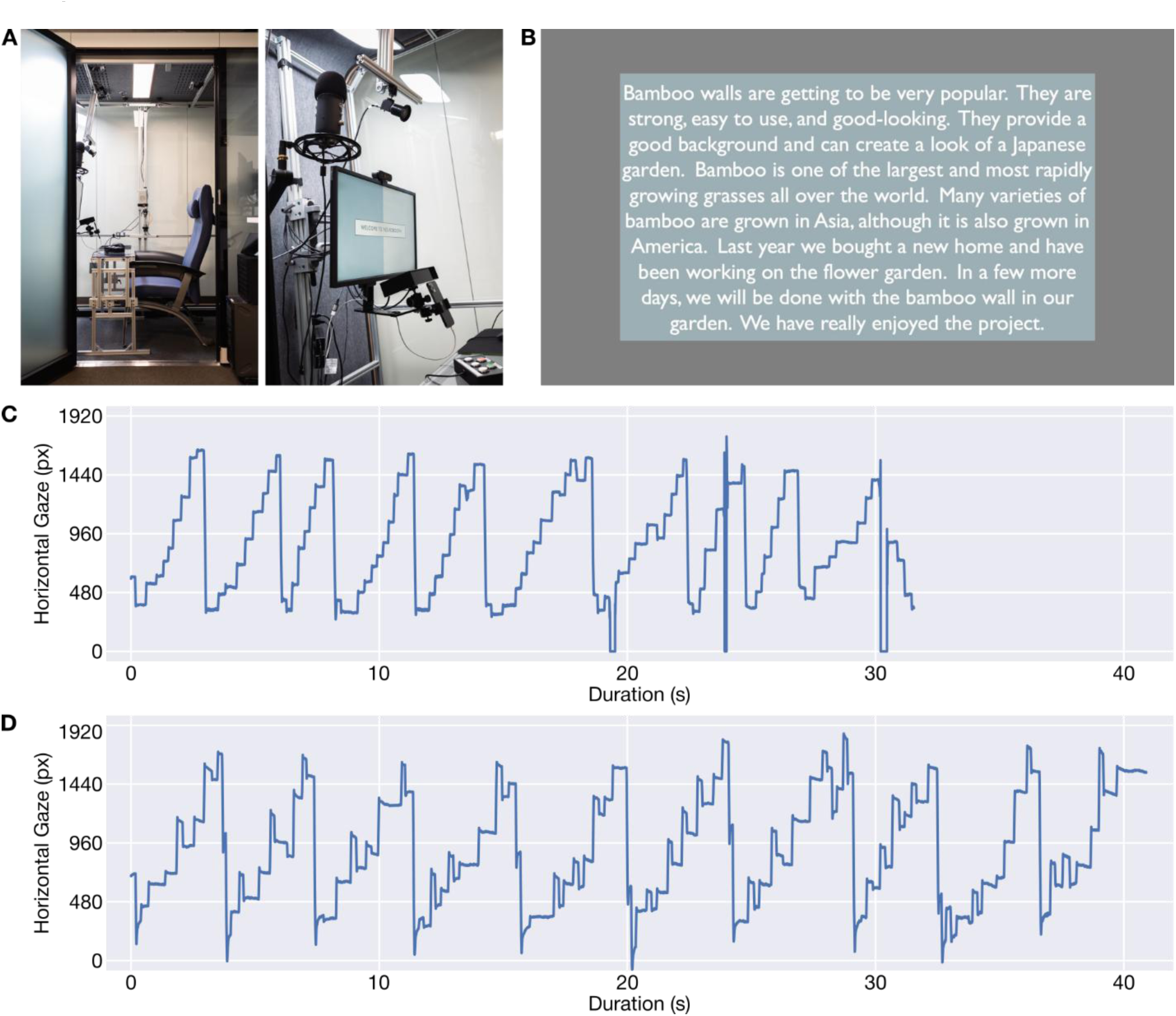
Experimental setup and data visualization. (**A**) The physical configuration of the Neurobooth, including the relative position of the chair, monitor, and EyeLink Portable Duo. (**B**) The Bamboo passage^23^ presented on the monitor to participants during the passage reading task. (**C**) The horizontal component of gaze during passage reading for a healthy control. (**D**) The horizontal component of gaze during passage reading for a participant with ataxia. The participant with ataxia took longer to read the passage and had more frequent regressions. Nystagmus, dysmetric saccades, saccadic intrusions, and saccadic pursuit were observed during clinical examination.

### Study Participants

Analyses were performed on data collected between April 28, 2022 and May 2, 2024. This data set included 139 individuals with clinically and/or genetically diagnosed ataxias and 78 healthy neurologically age-matched healthy controls. All participants provided informed consent and the research protocol was approved by the Massachusetts General Hospital Institutional Review Board (2021P000257, approved March 3, 2021).

A total of 45 participants (20.7%, 8 controls, 37 with ataxia) were subsequently excluded from analysis because of poor eye tracking data quality (see Supplementary Methods). Statistical comparisons were used to compare the clinical characteristics of included versus excluded participants (see Supplementary Methods). The demographics of the remaining 70 healthy controls and 102 participants with ataxia included in analyses are detailed in Table 1. There was no statistical difference (AUC=0.50, p=0.95) in the ages of the remaining healthy controls (54.9 ± 18.3 years old) and participants with ataxia (55.2 ± 16.5 years old). Longitudinal data were present for 37 controls and 26 individuals with ataxia. Consecutive visits were 85–623 (235.9 ± 93.2) days apart. The six most frequent diagnoses were Spinocerebellar Ataxia type 3 (SCA-3, N=19); Friedreich’s Ataxia (FRDA, N=12); SCA-6 (N=9); Cerebellar Ataxia, Neuropathy and Vestibular Areflexia Syndrome (CANVAS, N=7); SCA-2 (N=6); and the cerebellar subtype of Multiple System Atrophy (MSA-C, N=5).

**Table 1.** Participant demographics

| Diagnosis | *N* | Pct. Female | Pct. 4-Yr. Degree* | Decade of Life | BARS Total Mean (Range) | BARS Oculomotor Mean (Range) |
| --- | --- | --- | --- | --- | --- | --- |
| **Control** | **70** | **64** | **82** | **2nd–8th** | **—** | **—** |
| **Total Ataxia** | **102** | **47** | **52** | **2nd–8th** | **10.6**<br>**(0.0–25.0)** | **1.08**<br>**(0.0–2.0)** |
| Spinocerebellar Ataxia Type 1 | 2 | 100 | 0 | 3rd–6th | 13.5<br>(8.5–18.5) | 0.75<br>(0.5–1.0) |
| Spinocerebellar Ataxia Type 2 | 6 | 50 | 80 | 3rd–7th | 7.3<br>(0.5–14.0) | 0.83<br>(0.5–1.5) |
| Spinocerebellar Ataxia Type 3 | 19 | 63 | 44 | 2nd–7th | 9.3<br>(0.0–21.0) | 1.14<br>(0.0–2.0) |
| Spinocerebellar Ataxia Type 6 | 9 | 56 | 11 | 6th–8th | 12.2<br>(0.0–24.5) | 1.22<br>(0.0–2.0) |
| Friedreich's Ataxia | 12 | 33 | 50 | 2nd–7th | 18.1<br>(8.0–25.0) | 1.15<br>(0.0–2.0) |
| MSA-C | 5 | 60 | 67 | 6th–7th | 13.1<br>(9.0–18.5) | 1.00<br>(0.5–1.5) |
| CANVAS | 7 | 29 | 67 | 3rd–7th | 8.6<br>(0.0–23.0) | 1.21<br>(0.0–2.0) |
| HSP-7 | 4 | 25 | 50 | 4th–6th | 12.9<br>(7.5–16.0) | 0.88<br>(0.0–1.5) |
| Other Ataxias | 38 | 42 | 64 | 2nd–8th | 8.8<br>(0.5–18.0) | 1.07<br>(0.0–2.0) |

Abbreviations: BARS—Brief Ataxia Rating Scale; MSA-C—Multiple System Atrophy (Cerebellar Type); CANVAS—Cerebellar Ataxia, Neuropathy, and Vestibular Areflexia Syndrome; HSP-7—Hereditary Spastic Paraplegia Type 7
“Other Ataxias” included Spinocerebellar Ataxia Types 4, 7, 9, 17, 19, 22, 28, and 43; Gordon-Holmes syndrome; Behcet’s disease; autosomal recessive cerebellar ataxia; sensory ataxia; vestibular ataxia; pancerebellar syndrome; autoimmune-related ataxia with undefined cause; sporadic adult onset ataxia; and sporadic adult onset ataxia with neuropathy; and sporadic adult onset ataxia with downbeat nystagmus.
\* Percentage is computed with respect to the number of subjects who self-reported this information.

### Contextual Clinical Information

Participants with ataxia were assessed using the Brief Ataxia Rating Scale^3^ (BARS) during their clinical appointments, which has a total score range of 0–30 with 30 being the most severe. BARS subscores (either finger-nose or heel-shin) were missing for four visits corresponding to three participants. One participant was missing subscores worth eight points and the remaining participants were missing subscores worth four points. BARS total scores were normalized to account for missing subscore values. For the 102 participants with ataxia, normalized BARS ranged from 0–25 (10.6 ± 6.5) points. Fourteen participants had a BARS oculomotor score of 0 at the time of assessment, indicating no clinically-observed oculomotor signs during that assessment. (Three of these participants had increased BARS oculomotor scores on subsequent assessments.)

All participants were asked to complete a battery of patient-reported outcome measures (PROMs), including: PROM-Ataxia^24^; the depth perception, visual acuity and spatial vision, and visual processing speed sections of the Visual Activities Questionnaire^25^ (VAQ); the Dysarthria Impact Scale^26^ (DIS); the Communicative Participation Item Bank^27^ (CPIB); and ten of the thirteen Quality of Life in Neurological Disorders (NQoL) short forms^28^ (see Supplementary Methods).

### Data Processing

The time series from each eye were processed independently for every data recording session (see Supplementary Methods). If the task was repeated by the participant within a single session, then only the last repetition with a duration of at least 5 s was selected. Saccades were labeled as progressive (rightward) saccades, regressions (leftward), and sweeps (large displacement saccades primarily generated when scanning to the next line) based on their direction and displacement.

### Comparative Kinematic Measures

Saccade and fixation measures were computed using data from a single eye to facilitate comparison with the findings of Terao, et al.^20^ Kinematic measures included the mean angular displacement of saccades, the mean duration of saccades, the frequency of saccades, the mean duration of fixations, and the percentage of regressions. Saccade frequency was computed as the number of saccades divided by the cumulative duration of the saccades and fixations. In addition to these measures reported by Terao, et al., we also computed the number of saccades generated throughout the task recording and the mean angular displacement during fixations as the eyes are not entirely stationary during fixations.^29^ Sweeps were excluded from the calculation of all comparative kinematic measures.

Because passage reading is a multi-component task, we further examined the extent to which oculomotor and speech dysfunction, as measured by BARS, predicted each kinematic measure. Multiple regression was conducted for each kinematic measure with BARS speech and BARS oculomotor scores as predictor variables and the kinematic measure as the dependent variable. The relative contribution of each predictor variable to the model informed interpretation of how much dysfunction in each motor domain influenced passing reading performance.

### READ Score

Similarly to the comparative kinematic measures, a set of 28 kinematic data features were computed for each eye to summarize each task recording. Unlike the comparative measures, these features were extracted independently from the set of fixations and from the three saccade classes (i.e., progressive saccades, regressions, and sweeps). For fixations and each saccade class, six features were computed: the mean and coefficient of variation of duration, angular displacement, and peak angular speed. These features accounted for 24 of the 28 features. An additional feature for each saccade class was the percentage of saccades contained within the class, resulting in an additional three features. Finally, the saccade frequency feature was computed as the number of all saccades divided by the combined duration of all saccades and fixations.

The Reading Eye Abnormality Digital (READ) score was learned using Lasso regression to jointly model the relationship between the extracted features and total BARS (see Supplementary Methods). Total BARS, rather than the BARS oculomotor subscore, was used as the learning objective because 1) eye movements during passing reading in part reflect dysarthria^20^ and 2) not all cardinal oculomotor signs assessed the oculomotor component of BARS are necessarily present during reading.

### Statistical Analyses

Kinematic measures for participants with ataxia were grouped into low severity, mid severity, and high severity groups corresponding to total BARS of less than 8, 8–16, and greater than or equal to 16, respectively.^30^ Each measure was assessed based on its ability to differentiate between healthy controls and each of the severity groups. The READ score was assessed based on its correlation with established measures (BARS and PROMs), its ability to distinguish subclinical oculomotor signs, its reliability (agreement when independently assessing each eye), and its sensitivity to longitudinal changes. We also validated the READ score with respect to the amount of time each subject needed to read the passage, which is correlated with speech dysfunction. The Supplementary Methods describes the statistical analyses in further detail.

All statistical analyses used an a priori significance threshold of 0.05. Interpretations of effect size strength (i.e., weak, moderate, strong) for d and partial 𝜂^2^ were in accordance with guidelines proposed by Cohen.^31^ Effect sizes of 0.2, 0.5, and 0.8 were interpreted as weak, moderate, and strong for Cohen’s d, respectively. Effect sizes of 0.01, 0.06, and 0.14 were interpreted as weak, moderate, and strong for partial 𝜂^2^, respectively.

## Results

### Comparative Kinematic Measures

Several saccade and fixation kinematic measures during reading were observed to change with ataxia severity and differ between ataxia and control groups (Fig. 2, Supplementary Table 1). In particular, the angular displacement of detected fixations was strongly related to overall ataxia severity group (𝜂^2^=0.44, p<0.001). Severity was also strongly associated with an increased number of saccades (𝜂^2^=0.27, p<0.001) and reduced saccade frequency (𝜂^2^=0.25, p<0.001), and was moderately associated with an increased percentage of regressions (𝜂^2^=0.11, p<0.001). In addition to reduced saccade frequency, a moderate association was observed between severity and fixation duration (𝜂^2^=0.13, p<0.001). Notably, both the angular displacement of fixations (d=0.72, p=0.002) and the number of saccades (d=0.58, p=0.02) were different between controls and low-severity ataxia.

**Figure 2.**
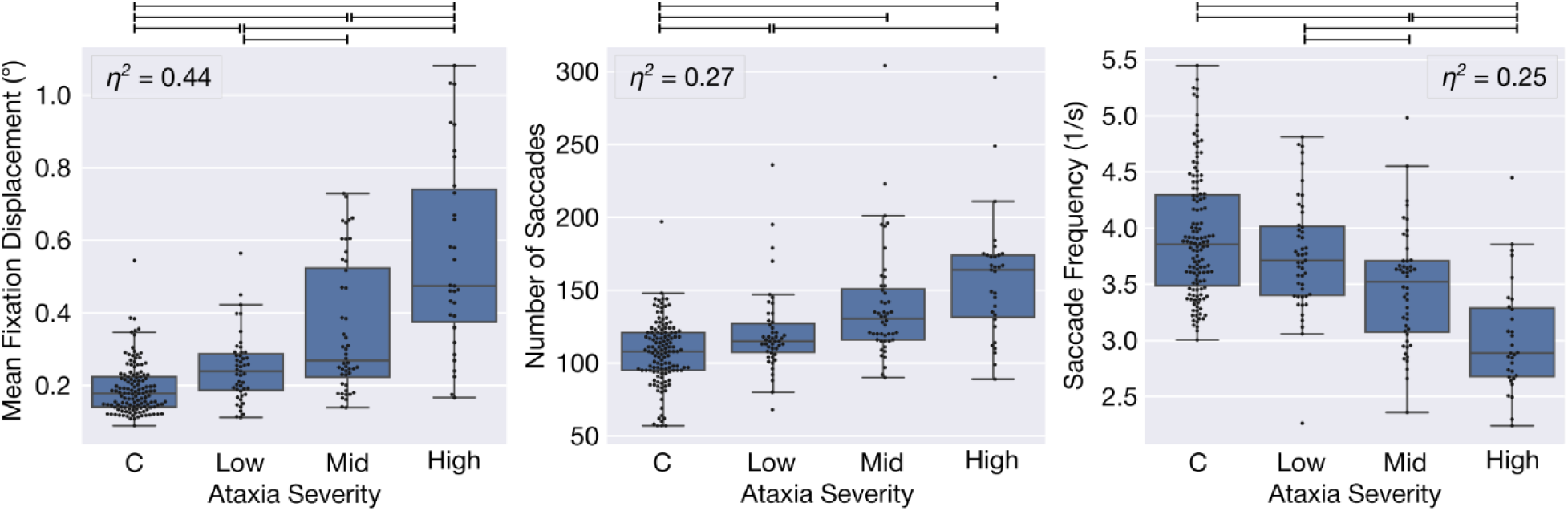
Comparative kinematic measures of interest. Sweeps were excluded from measure calculations. Participants were grouped into controls (C), low severity, mid severity, and high severity based on the total Brief Ataxia Rating Scale. Box plots denote each quartile, with a maximum whisker length of 1.5 times the interquartile range. The overlaid swarm shows each task recording. The horizontal black bars above each plot denote statistical significance of the post-hoc tests comparing the indicated groups. The omnibus test was not significant for mean saccade displacement. For other measures, *p*-values were less than 0.001.

To evaluate whether eye movement kinematic measures were more closely related to eye movement dysfunction or speech dysfunction, we computed the relative percentage of variance of each measure explained by BARS speech and oculomotor subscores (Table 2). Changes in fixation displacement were best explained by changes in oculomotor dysfunction. Changes in the number and frequency of saccades and in fixation duration were instead better explained by changes in speech dysfunction. Therefore, both speech and oculomotor dysfunction were reflected by changes in the kinematics of saccades and fixations during passage reading.

**Table 2.**
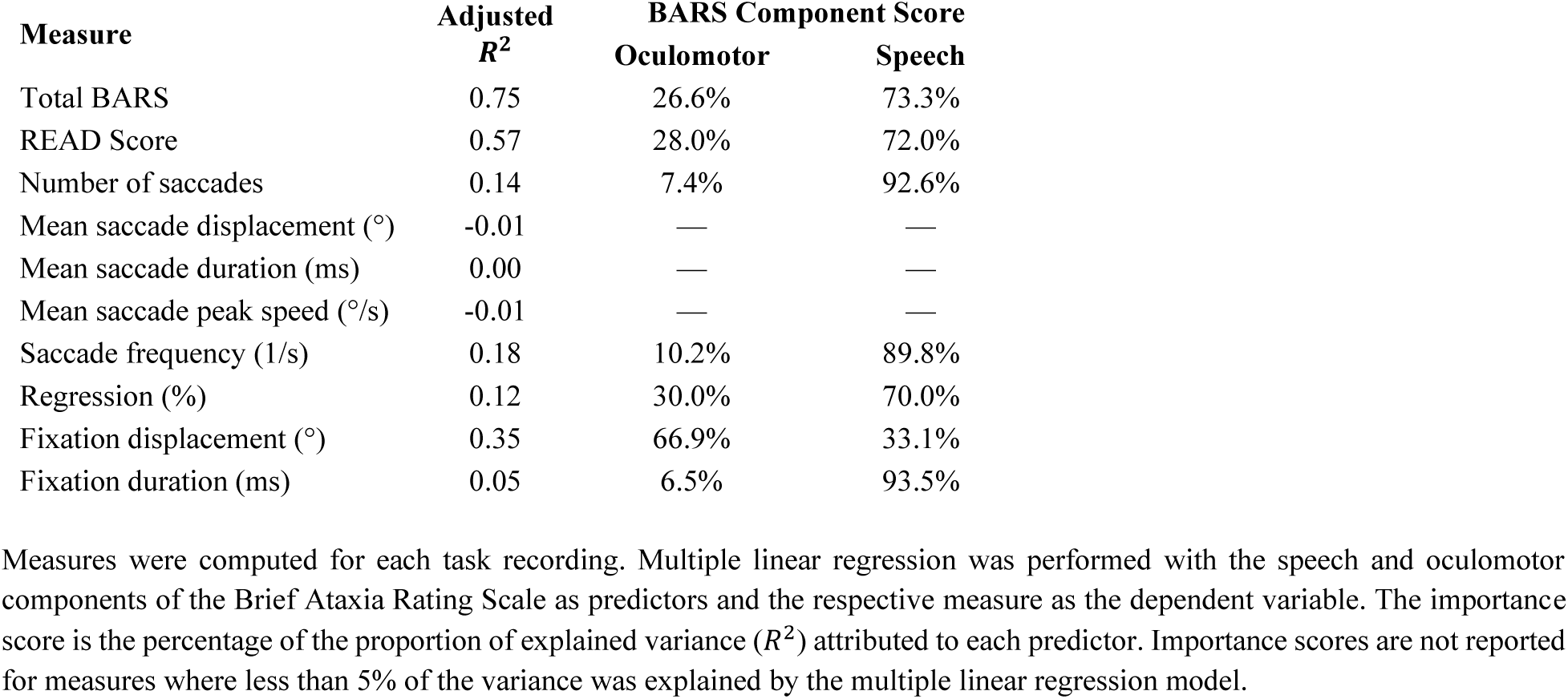
Amount of variance explained by oculomotor and speech dysfunction

### READ Score Evaluation

Next, we evaluated the properties of the machine-learned composite measure (READ score) that was trained to take a weighted sum of all the eye movement kinematic features in order to estimate the BARS total score. The READ score most heavily weighted the percentage of sweeps, fixation displacement, saccade frequency, fixation duration variability, and the percentage of regressions (see Supplementary Table 2 and Supplementary Results).

#### Reliability and relationship with overall ataxia severity

The READ score was reliable (ICC=0.96, p<0.001), had a strong correlation with total BARS (r=0.82, p<0.001), and accurately distinguished between controls and participants with ataxia (AUC=0.89, p<0.001, Fig. 3). The READ score association with total BARS was consistent across most specific diagnoses (Supplementary Fig. 6, Supplementary Results).

**Figure 3.**
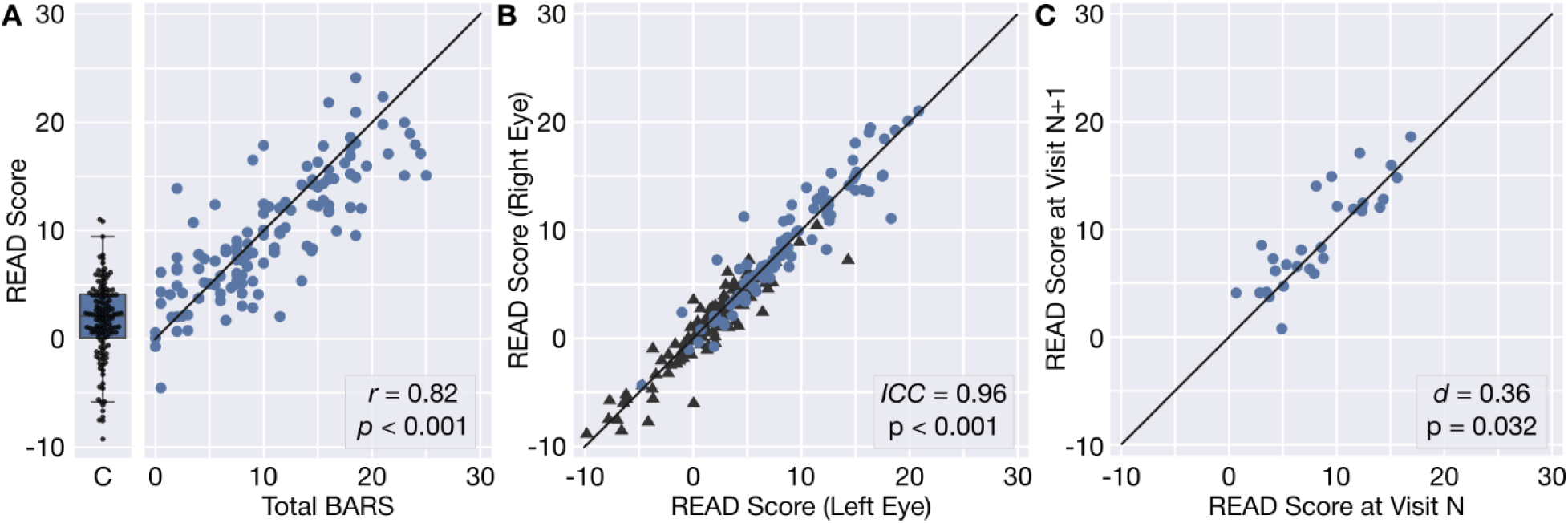
Per-recording validation and characterization of the READ score. In all plots, the solid black line denotes perfect agreement and markers correspond to a single task recording. (**A**) The READ score (averaged across both eyes) strongly correlated with total Brief Ataxia Rating Scale (BARS). The distribution of the READ score is shown separately as a swarm for healthy controls, as BARS was not assessed. The box plot denotes each quartile, with a maximum whisker length of 1.5 times the interquartile range. (**B**) The READ scores independently obtained from each eye from the same task recording demonstrated very strong agreement. Healthy controls are denoted by black triangles. (**C**) The READ scores from consecutive visits (at least 85 days apart) demonstrated sensitivity to longitudinal change; validated by a one-sided, one-sample *t*-test.

#### Sensitivity to change

The READ scores from consecutive study visits for participants with longitudinal data were compared to determine if the score increased over time, consistent with disease progression over time. Indeed, the READ score demonstrated sensitivity to longitudinal change across multiple visits (d=0.36, p=0.03) for participants with ataxia. Change in the READ score over time was not observed in healthy controls (d=0.09, p=0.25).

In contrast, total BARS (d=0.27, p=0.08), BARS speech (d=0.30, p=0.06), and BARS oculomotor (d=0.09, p=0.31) scores did not reach statistical significance in measuring longitudinal change in the study cohort.

#### Sensitivity to subclinical signs

The READ score’s ability to distinguish between controls and participants with ataxia with a BARS component score of zero was evaluated to determine whether the READ score was sensitive to subclinical signs of ataxia. This evaluation was independently performed for each BARS component score. The READ Score demonstrated sensitivity to subclinical signs (i.e., a score of zero) for the oculomotor (AUC=0.69, p=0.02), speech (AUC=0.72, p<0.001), and finger-nose (AUC=0.69, p=0.01) components of BARS. No sensitivity was observed with respect to the heel-shin (AUC=0.58, p=0.34) and gait (AUC=0.53, p=0.78) components of BARS. The significance for the finger-nose component may be in part due to its strong relationship with the remaining components of total BARS (Supplementary Fig. 7).

#### Association with patient-reported measures of function

The READ score was moderately correlated with PROMs that captured motor and speech symptoms of ataxia but not with PROMs capturing affect, cognition, or visual deficits (Table 3). Correlated PROMs capturing motor symptoms included the PROM-Ataxia total score (r=0.51, p<0.001), the PHYS-2 (physical capability; r=0.62, p<0.001) and ADL (activities of daily living; r=0.56; p<0.001) components of PROM-Ataxia, and the NQoL upper extremity (fine motor capabilities; r=-0.47; p<0.001) and lower extremity (mobility; r=-0.53, p<0.001) short forms. The PHYS-1 component of PROM-Ataxia, which includes questions relating to autonomic, sensory, and sleep dysfunction, was only weakly correlated with the READ score (r=0.37, p<0.001). The READ score also correlated with PROMs capturing speech symptoms, including the Communicative Participation Item Bank (r=-0.54, p<0.001) and Dysarthria Impact Scale (r=0.53, p<0.001). The READ score was weakly correlated with visual processing speed (r=0.27, p=0.01) and not correlated with depth perception (r=0.17, p=0.10) or visual acuity (r=0.00, p=0.99), as measured by the Visual Activities Questionnaire (see Supplementary Results).

**Table 3.** Correlations of contextual clinical measures with learned composite scores.

| PROM | PROM Component | Correlation with Composite Score |  |
| --- | --- | --- | --- |
| | | $r$ | $p$ |
| PROM-Ataxia | (total) | 0.51 | <0.001 |
|  | Physical Section 1 | 0.37 | <0.001 |
|  | Physical Section 2 | 0.62 | <0.001 |
|  | Activities of Daily Living | 0.56 | <0.001 |
|  | Mental Section 1 | 0.14 | 0.13 |
|  | Mental Section 2 | 0.35 | <0.001 |
| Visual Activities Questionnaire | Depth Perception | 0.17 | 0.10 |
|  | Visual Processing Speed | 0.27 | 0.01 |
|  | Visual Acuity and Spatial Vision | 0.00 | 0.99 |
| Dysarthria Impact Scale | (total) | 0.53 | <0.001 |
| Communicative Participation Item Bank | (total) | -0.54 | <0.001 |
| Quality of Life in Neurological Disorders (short forms) | Lower Extremity | -0.53 | <0.001 |
|  | Upper Extremity | -0.47 | <0.001 |
|  | Participation in Social Roles | -0.34 | <0.001 |
|  | Wellbeing | -0.22 | 0.02 |
|  | Cognition | -0.13 | 0.17 |
|  | Fatigue | -0.01 | 0.95 |
|  | Sleep | 0.00 | 0.97 |
|  | Depression | 0.13 | 0.18 |
|  | Anxiety | 0.05 | 0.57 |
|  | Emotional Dyscontrol | 0.15 | 0.12 |

#### Relationship with clinician-assessed domain severity

We further investigated the relationship between the READ score and the BARS component scores (Fig. 4). The READ score was moderately correlated with BARS oculomotor (r=0.52, p<0.001) and strongly correlated with BARS speech (r=0.73, p<0.001). The READ score was also correlated with BARS finger-nose (r=0.72, p<0.001), heel-shin (r=0.67, p<0.001), and gait (r=0.73, p<0.001).

**Figure 4.**
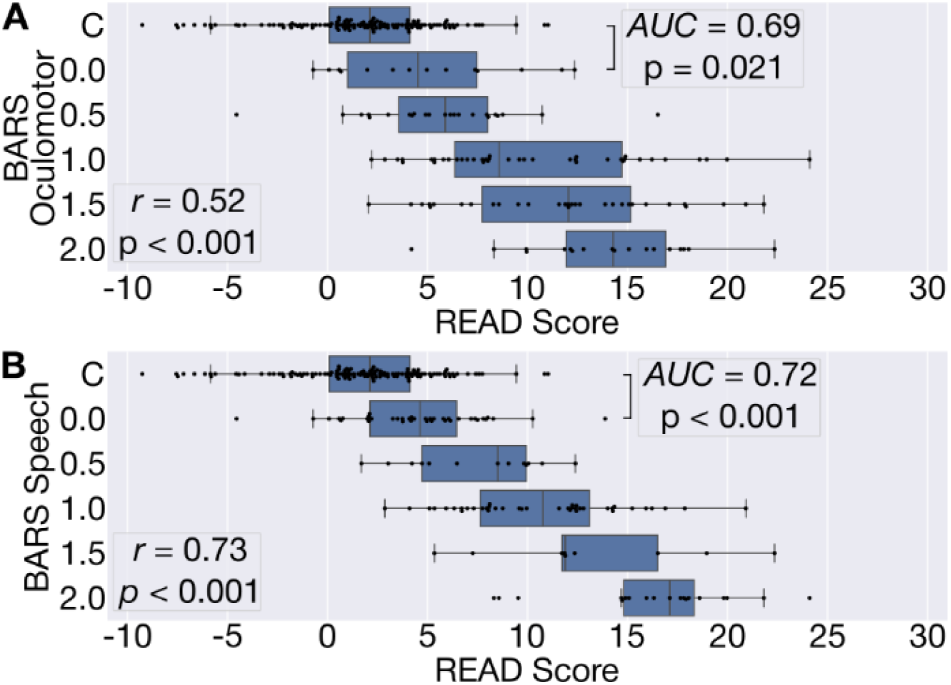
READ score versus (A) BARS Oculomotor and (B) BARS Speech. The READ score increased with respect to BARS subscores and demonstrated some sensitivity to subclinical signs. No participant with ataxia had a BARS Speech score of more than 2. Box plots denote each quartile, with a maximum whisker length of 1.5 times the interquartile range. The overlaid swarm shows each task recording. READ score differences between controls and participants with ataxia with a BARS subscore of zero were significant.

#### READ score as a function of participant severity, age, sex, and education

We investigated the relationship between the READ score and total BARS, age, sex, attainment of a four-year university degree, and disease status (i.e., ataxia vs. healthy control) using multiple linear regression. A total of 66.9% of the variance in the READ score was explained by these factors. The relative percentage of the explained variance was 60.3% for total BARS, 25.1% for disease status, 8.8% for attainment of a four-year university degree, 4.7% for sex, and 1.4% for age.

#### Comparison with passage reading task duration

Given the relationship between the READ score and speech dysfunction, we investigated whether the READ score was more informative than the time needed to complete the passage reading task (i.e., an easily measured proxy for speech dysfunction). The reading duration was able to separate the severity groups (𝜂^2^=0.45, p<0.001) and controls from participants in the low severity group (d=-0.66, p<0.001). However, the READ score was more strongly correlated with total BARS (r=0.82, p<0.001, 95% CI: 0.75–0.87) than reading duration (r=0.61, p<0.001, 95% CI: 0.48–0.71). The READ score also better captured longitudinal changes (d=0.36, p=0.03) than reading duration (d=0.24, p=0.10). In contrast, reading duration was similarly sensitive to subclinical signs of oculomotor (AUC=0.73, p=0.005) and speech dysfunction (AUC=0.68, p=0.001) as the READ score. Multiple regression with total BARS, age, sex, attainment of a four-year university degree, and disease status as predictors explained 55.1% of the variation in reading duration. The relative percentage of the explained variance was 53.2% for total BARS, 19.4% for disease status, 18.3% for attainment of a four-year university degree, 8.3% for sex, and 0.8% for age.

## Discussion

We found that eye movement kinematics during reading—including mean displacement of fixations, the number and frequency of saccades, and the percentage of regressions—were related to speech severity, oculomotor severity, and overall severity in a large ataxia cohort. By quantitatively aggregating saccade and fixation properties, we developed the READ composite score which demonstrated key measurement properties. The READ score had high reliability, captured early speech and oculomotor changes, and was more responsive to disease progression than clinical assessments.

Our finding that the kinematics of eye movements while reading aloud are informative of the presence and severity of ataxia is supported by prior studies. The mean displacement of fixations captures fixational instability, which is a hallmark of the ataxia phenotype.^32–35^ Oh, et al. also observed greater fixation dispersion during regularly-spaced number reading.^21^ Furthermore, Terao, et al. reported an increased percentage of regression saccades in participants with ataxia when reading Japanese text aloud.^20^ This increased percentage of regressions likely contributed to our observation of both an increased number of progressive saccades^21^ and reduced percentage of large sweep saccades (due to the increased number of regressive and progressive saccades per line). This increased percentage of regressions and larger number of total saccades may contribute to additional concentration and fatigue during reading and other tasks requiring sequential control of eye movements.^21^ We also observed a strong negative relationship between saccade frequency (i.e., saccades per second) and ataxia severity that was not observed by Terao, et al.^20^ Our identification of this signal is likely due to the larger size of our cohort and the use of a more fine-grained proxy for severity (i.e., BARS) than disease stage. Additional factors, including the language of the reading task (Japanese is logographic and has a significant vertical component) may have also contributed to this finding. Reduced saccade frequency is supported by studies that reported slower reading speeds and increased fixation duration during silent reading.^21,35^ Interestingly, we did not observe significant differences in the displacement, peak speed, or duration of progressive saccades. The lack of difference in mean saccade displacements may be due to the small and variable size of saccades generated during passage reading. The lack of difference in mean saccade durations or peak speeds may additionally be due to the heterogeneity of the diagnoses included in this study, as not all ataxias are associated with saccadic slowing.

The majority of the variance in the READ score was explained by the presence and severity of ataxia. Contributions from sex and age were minimal (together explaining less than 4% of the READ score’s variance). However, we observed a small contribution related to participants’ level of education. This effect may be due to a likely association between educational attainment and reading skill, which is known to affect eye movement kinematics during reading. In addition to perceiving an increased amount of text while fixating, greater reading skill is associated with shorter fixations, longer saccades, and fewer regressions.^29^ Therefore, educational level should likely be controlled for in multi-site studies performing group comparisons where large differences in education demographics are likely. It is also interesting that the variance in reading duration was much less explained by ataxia presence and severity and more explained by educational level. This finding further supports the READ score’s ability to more sensitivity capture disease severity and progression than the simpler measure of task duration.

Though reading aloud is a multi-component task, the current analysis focused on eye movements. However, we found that the READ score and several eye movement abnormalities more strongly reflected speech dysfunction than oculomotor dysfunction with respect to both clinician-scored and patient-reported outcome measures. These findings are likely due to the complex interactions between dysarthric speech and eye movements while reading aloud.^20^ For example, eye movements are slowed while reading aloud so that progression of gaze along the text does not outpace speech production.^20,29^ While eye movements during reading capture information about speech, they do not necessarily reflect all the cardinal oculomotor signs of ataxia. In particular, pursuit movements are unlikely to be generated during reading. Therefore, analysis of either pure oculomotor tasks^15^ or additional functionally relevant tasks that elicit both smooth pursuit deficits and many large, potentially dysmetric saccades (e.g., watching a carefully-chosen video) may be necessary to fully capture the full spectrum of oculomotor dysfunction caused by cerebellar ataxias.

Complex, multi-component behaviors remain a rich and underexplored potential source of novel digital measures of neurologic disease. The READ score’s sensitivity to disease progression and ability to capture early disease signs demonstrates this potential, and supports incorporation of quantitative oculomotor reading assessments into clinical research in ataxias^21^. This study also highlights the possibility of inclusion in routine clinical practice in the future, given that the eye tracking data were collected at scale in an active clinical setting. Furthermore, because reading is fundamental to many aspects of everyday modern life, it is a promising target as a common, meaningful, and structured behavior to capture and assess. Ataxia trials considering the use of other functionally-relevant digital endpoints, such as wearable- or smartphone-based gait biomarkers^36,37^, may include reading-based measures as a complementary endpoint. In the near future, advances in the use of mobile phone cameras for eye tracking,^36,38–40^ in conjunction with innovations in smart glasses and extended reality technologies, may also provide the technical capability to enable high quality recording of eye movements at home and at massive scale^41^. However, additional research is needed to validate the usability of kinematic measures obtained from these devices in realistic settings given the potential impacts of varied environmental conditions and slippage^42^ on data quality.

The strong measurement characterics of the READ score motivates future inquiry into the quantitative assessment of reading in neurologic disease populations. Firstly, reading presents an opportunity to capture signs of cognitive deficits in addition to speech and oculomotor dysfunction.^29,43^ Second, it is likely that additional quantitative modeling techniques, such directly modeling progression^44^ or loss functions that do not encourage absolute agreement with clinical scales,^45,46^ can further improve the score’s sensitivity for measuring disease progression. Given the rich temporal structure of eye movements during reading, modeling strategies that can effectively leverage the temporal structure may also yield more sensitive measures. Finally, additional sensitivity is likely to be gained from the integration of both quantitative speech analyses^14,18,47^ and analysis of the interaction between speech and eye movements.^20^

There were some limitations to the study. Data were collected as part of a larger study in a clinic setting under time constraints, which may have contributed to the need to exclude some sessions due to poor calibration. These exclusions resulted in a slight inclusion bias towards individuals with milder disease severity, although more severe individuals were well represented in the study. Including real time feedback on data quality and allowing additional time for setup and calibration (and repetition of the task when needed) could substantially reduce the percentage of sessions with poor data quality. Additional time would also permit reading of multiple passages that would provide additional data to further reduce the likelihood of exclusion. It is also possible that some controls included in the study were not true controls, as we relied on self-reported medical history rather than a detailed neurological assessment of controls. However, the READ score was able to distinguish between controls and participants with ataxia despite this possibility. Only the horizontal component of gaze was analyzed in this study. Reading in other languages has a more significant vertical component, which may complicate study replication in many geographic regions.

The READ score highlights the potential for sensitive, functionally relevant measures of eye movements in ataxias and other neurodegenerative diseases affecting speech or oculomotor control. Though we demonstrate practicality in an active clinical setting using specialized hardware, the advancement of at-home eye tracking technologies may further extend the scalability of this assessment approach.

## Data Availability

Data can be requested by qualified researchers by visiting https://neurobooth.mgh.harvard.edu/.

## Acknowledgements

This project utilized resources at the Martinos-MLSC cluster. The authors would like to thank prior contributors to the Neurobooth project not directly involved with this research, including Adonay Nunes, Mainak Jas, Nicole Eklund, and Sheraz Khan. The authors also thank the Neurobooth participants for their valuable time and perspectives.

## Supplementary Methods

### A. Data Collection

The Neurobooth is situated in the neurology clinic at Massachusetts General Hospital (MGH) and supports time-synchronized data collection using a large array of sensors to capture performance on a variety of behavioral stimuli. The task battery for the Neurobooth study is designed to be completed within 30-40 minutes. Eye tracking calibration was performed using a five-point stimulus at the beginning of the task battery. In some situations, the calibration was not able to be validated due to severe disease signs (e.g., saccadic intrusions). After calibration, participants completed a battery of oculomotor (e.g., saccades, smooth pursuit, fixation) and speech (e.g., sustained phonation, repeated consonants) tasks before being presented with the passage reading task. Participants took as much time as needed to read the passage aloud. A research coordinator concluded the recording when the participant was finished reading.

Stimuli were presented on a 24-inch BenQ monitor with a resolution of 1920 by 1080 pixels and a refresh rate of 240 Hz. The monitor height was adjustable, though to maintain throughput on busy days it was only adjusted (such that the top of the monitor was at eye level) for particularly tall or short participants. An EyeLink Portable Duo (SR Research Ltd.) was mounted on a bracket attached to the monitor mount such that the center of the eye tracker was 12 cm in front of and vertically level with the bottom of the screen. A target sticker with concentric black and white circles was affixed to the center of participants’ foreheads during data collection.

### B. Initial Exclusion of Poor-Quality Data

The quality of each task recording was assessed using three criteria. The first criteria was designed to reject recordings with poor calibration. Every sample where the horizontal or vertical gaze position exceeded the known boundaries of the screen by at least 20% were flagged as out-of-bounds. The second criteria was designed to reject recordings with excessive missing data. Every sample that had a zero (i.e., placeholder) value or that was labeled as a blink by the eye tracker was flagged as being missing data. If at least 20% of samples in the task recording were flagged as either out-of-bounds or missing, then the task recording was excluded from analysis. The third criteria was designed to reject recordings where the participant’s head excessively moved. If the standard deviation of the distance between the forehead marker and the eye tracker during the recording exceeded 100 mm, then the recording was excluded from analysis. After data quality exclusions, the mean, 95th percentile, and maximum of the standard deviation of the forehead marker distance in each recording was 3.6 mm, 10.7 mm, and 31.1 mm, respectively.

### C. Patient-Reported Outcome Measures

Patient-reported outcome measures (PROMs) were administered remotely using REDCap. The PROM-Ataxia consists of 70 items scored on a 0–4 Likert scale distributed across five components assessing affect (MEN-1), cognition (MEN-2), activities of daily living (ADL), physical symptoms including sleep, balance, and sensation (PHYS-1), and physical capabilities (PHYS-2). The DIS (23 items) and CPIB (10 items) assess how speech impairments affect everyday life and how patients’ conditions impact how they communicate on an average day, respectively. The NQoL short forms, each consisting of 8–9 items, assess the ability to perform fine motor tasks, mobility difficulties, cognition, anxiety, depression, life satisfaction, emotional control, fatigue, sleep disturbances, and patients’ abilities to keep up with social responsibilities.

Prior to May 16, 2023, there was a REDCap error in the first mental (affect) section of PROM-Ataxia that combined the “rarely” and “sometimes” responses (with respective scores of 2 and 3) into a single response. A value of 2.5 was used for responses in this erroneous category.

### D. Data Processing

Time series were automatically trimmed to the performance of the passage reading task using timestamps logged by the Neurobooth software (Figs. 1C and 1D). Saccades and blinks were detected using the eye tracker’s built-in algorithms. Fixations or saccades within 100 ms of a blink were excluded from analysis.^1^ Detected saccades with a duration of less than 6 ms were ignored (i.e., treated as part of the adjacent fixations).^2,3^ The vertical component of gaze was not considered in this analysis. Horizontal gaze angular velocities (°/s) and displacements (°) were derived as detailed below and in the EyeLink Portable Duo User Manual^1^.

#### I. Computation of Horizontal Angular Gaze Velocity

Let 𝑥(𝑡) and 𝑟_𝑥_(𝑡) be the horizontal gaze position (px) and horizontal angular resolution (px/°) at time 𝑡, respectively. The derivative of the horizontal gaze position with respect to 𝑡, 𝑥′(𝑡), was numerically computed using a second-order central differences method. The angular horizontal gaze velocity (°/s) was computed as 𝑥′(𝑡) / 𝑟_𝑥_(𝑡). Angular velocities were filtered using a low-pass, seventh-order Butterworth filter with a cutoff frequency of 100 Hz.^4^

#### II. Computation of Horizontal Angular Gaze Displacement

Given two horizontal gaze coordinates in the head-referenced (HREF) plane, 𝑥_1_ and 𝑥_2_, the horizontal displacement in degrees visual angle was computed as,

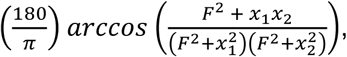

where 𝐹 = 15,000 is a constant specifying the distance of the HREF plane from the eye. (This unit is independent of system setup, display distance, and display resolution.^1^) Saccades with an angular displacement less than 0.5° were excluded from subsequent analysis.^3^

#### III. Classification of Saccade Types

Each saccade was then classified as a progressive saccade, regression, or sweep based on its displacement and direction. Saccades with an angular displacement of at least 15° were classified as “sweeps”. This threshold was empirically determined based on the distribution of saccade angular displacements (Supplementary Fig. 1). Sweeps included both return sweeps (movements to the next line) and regressive sweeps (returning to the previous line). Non-sweep saccades were classified as regressions if participant gaze moved in a leftward direction on the screen. Otherwise, the saccade was classified as a progressive saccade.

#### IV. Exclusion of Data with Insufficient Saccades

Additional data quality filters related to the performance of the passage reading task were applied to ensure saccade-based data features could be reliability computed. More specifically, task recordings with less than forty saccades or less than four sweeps were excluded from further processing and analysis, although all task recordings with less than forty saccades also had less than four sweeps (Supplementary Fig. 2 and Supplementary Fig. 3).

#### V. Eye Selection for Comparative Kinematic Measures

Because Terao, et al.,^5^ used monocular recordings, we selected a single eye from each binocular recording for the computation of comparative kinematic measure. The eye ipsilateral to the dominant upper limb was used if available.^6–8^ If the data from the ipsilateral eye was unavailable or excluded as having poor quality, then the contralateral eye was used.

### E. Excluded Participants

The BARS total score for excluded participants with ataxia ranged from 2.5–23.5 (13.4 ± 5.3; mean ± standard deviation) points. BARS total ranged from 0–25 (10.6 ± 6.5) points for included participants, reflecting a large overlap. However, the difference in average overall disease severity was statistically significant (d=0.49, p=0.008). Excluded subjects also had higher BARS oculomotor scores (d=0.45, p=0.007). The 𝜒^2^ test for independence was used to evaluate differences in the presence of saccadic intrusions and in the presence of nystagmus between included and excluded participants. Effect sizes were assessed using 𝜑. The presence of saccadic intrusions was only weakly associated with exclusion from the study (𝜑=0.26, p=0.005). The presence of nystagmus was not associated with exclusion from the study (𝜑=0.05, p=0.600). Of the 45 participants excluded from the study, only seven exclusions corresponded to the minimum saccade and sweep criteria. Six out of seven of these participants had ataxia and all six had a BARS oculomotor score of at least 1.0.

### F. Stratification of Controls

Controls were classified as MGH controls or general population controls based on the recruitment mechanism. MGH controls were the family and caregivers of patients who received care at Massachusetts General Hospital Department of Neurology. General population controls were recruited more broadly using flyers, the Neurobooth website, and advertisements on the Rally for Partners platform. The distinction between controls was made because recruitment from the general population may result in inclusion of some individuals who participate due to a concern about their neurological function.^9–11^

### G. READ Score

Lasso regression was used to jointly model the relationship between the extracted features and total BARS. Features extracted from each eye recording were treated as independent model inputs, and the two model outputs were averaged. Though they were not scored on BARS, controls were assumed to have a total BARS of 0.0 for the purposes of model training. Only the MGH controls were included in the training data to reduce the potential for noise introduced by this assumption, although models were evaluated against all controls. Models were trained and evaluated using leave-one-subject-out cross-validation (LOSOCV). Model outputs corresponding to the testing set of each iteration of LOSOCV were pooled to facilitate model evaluation and statistical analyses.

Models were trained and evaluated using leave-one-subject-out cross-validation (LOSOCV). In each iteration of LOSOCV, one subject’s data (i.e., all task recordings across longitudinal visits) were withheld as the testing set and remaining subjects’ data were used as the training set. Feature standardization and hyperparameter selection (including random sampling of candidate regularization strengths) were independently performed within each iteration of LOSOCV. This procedure aims to mimic the generalization of a trained model to a previously unseen subject’s data.

For each iteration of LOSOCV, all features were standardized such that each feature in the training set had zero mean and unit variance. Then, 100 candidate values of the regularization strength hyperparameter of Lasso were randomly sampled from the log-uniform distribution spanning 10^−3^–10^3^. The hyperparameter maximizing the coefficient of determination (assessed using five-fold cross validation within the training set) was used when training each model.

### H. Statistical Analyses

The mean and standard deviation amongst controls and participants with ataxia was computed for each comparative kinematic measure. Welch’s t-tests were used to determine if the mean of each measure was significantly different between controls and participants with ataxia. Effect sizes were assessed using Cohen’s d. Welch’s ANOVA with Games-Howell post hoc tests were used to determine if there were significant differences in kinematic measures between controls and the three severity groups (based on total BARS). Effect sizes for Welch’s ANOVA and Games-Howell post hoc tests were assessed using partial 𝜂^2^ and Cohen’s d, respectively. The post-hoc tests comparing healthy controls to participants with low-severity ataxia were of particular interest.

Pearson’s correlation (r) was used to assess the convergent validity of the READ score with total BARS, each component score of BARS, and each PROM included in the study. The ability of the READ score to distinguish between participants with ataxia and controls was assessed using a Mann-Whitney U-test. Similar tests were performed for participants with ataxia and BARS oculomotor or speech scores of 0 to assess the READ score’s ability to distinguish subclinical oculomotor signs. Effect sizes were assessed in terms of the common language effect size, which is equivalent to the area under the receiver operating characteristic curve (AUC). The correlation between participant age and the absolute error of the READ scores (with respect to total BARS) was used to determine if the accuracy of the READ score was significantly influenced by participant age. READ score reliability was assessed by comparing the READ score for each eye during the same task recording using a two-way mixed effects, absolute agreement, single rater intraclass correlation coefficient (ICC).^12^ The sensitivity of the READ score to longitudinal changes was assessed using a one-sided, one-sample t-test to determine if the mean difference in READ scores between consecutive visits was greater than zero. The effect size was assessed using Cohen’s d. Analysis of the extent to which the kinematic measures and the READ score were explained by BARS component scores (Table 2 and Supplementary Table 3) were based on a linear regression model. The proportion of explained variance was measured using the adjusted coefficient of determination (R^2^) and relative importance was measured by averaging over all combinations of predictors.^13^

## Supplementary Results

### A. Comparative Kinematic Measures

As expected, the mild, moderate, and high severity groups (defined by BARS total score) were in general agreement with BARS oculomotor (𝜂^2^=0.24, p<0.001) and BARS speech (𝜂^2^=0.64, p<0.001) subscores (Supplemental Fig. 4).

Neither saccade displacement (𝜂^2^=0.03, p=0.09) nor duration (𝜂^2^=0.01, p=0.35) were found to be significantly different with respect to ataxia severity or between ataxia and control groups (Supplementary Table 1). We therefore hypothesized that the increased number of saccades in individuals with ataxia was due to the increased percentage of regression saccades that result in increased visual traversal of the passage. Indeed, the number of progressive and regressive saccades were correlated (r=0.55, p<0.001) and were both strongly indicative of disease severity (Supplementary Fig. 5). We also observed a strong correlation between the cumulative displacements of progressive and regressive saccades (r=0.83, p<0.001), supporting the hypothesis of increased visual traversal. It is therefore notable that the number of saccades provided a stronger signal than the percentage of regressions for both ataxia severity and for separating ataxia and control populations.

### B. Feature Importance

The severity estimation model was probed to understand the relative contribution of each saccade and fixation feature to the READ score. Higher READ scores (indicating increased disease severity) were predominantly driven by a decreased percentage of sweeps, increased fixation displacement, decreased saccade frequency, increased variability of fixation durations, and an increased percentage of regressions (Supplementary Table 2). These feature importances are consistent with the strong trends observed in the kinematic measures of saccades and fixations (Fig. 2 and Table 2).

### C. READ Score for Controls

There was no difference in the READ score of general population controls and MGH controls (AUC=0.52, p=0.76).

### D. READ Score for Frequent Specific Diagnoses

Because the study population included a diverse range of ataxias, we evaluated the relationship between the READ score and BARS total separately for the six most-frequent ataxia diagnoses in the study (Supplementary Fig. 6). The four most-frequent diagnoses (SCA-3, FRDA, SCA-6, and CANVAS) all demonstrated similar trends to the READ score across all diagnoses. The READ score for MSA-C and SCA-2 exhibited some differences with respect to overall READ score. The sample size was smallest for MSA-C and SCA-2 (N=5 and N=6, respectively) and participants spanned a narrow range of disease severity (Supplementary Fig. 6). For SCA-2, the observed difference in agreement may be in part due to saccadic slowing not being well captured by the reading task. Though we observed a statistical difference in mean peak saccade speed between controls and individuals with SCA-2 (d=0.69, p=0.002), these observed mean speeds did not correlate with total BARS (r=0.05, p=0.91).

### E. READ Score Relation to Speech and Oculomotor Dysfunction

A total of 57% of the variance in the READ Score was explained by the BARS oculomotor and BARS speech scores, with 28.0% of the explained variance attributed to BARS oculomotor and the remaining 72.0% attributed to BARS speech (Table 2). In comparison, 75% of the variance in total BARS was explained by BARS oculomotor and BARS speech scores, with similar attributions to BARS oculomotor (26.6%) and BARS speech (73.3%).

When considering all BARS component scores, a total of 67% of the variance in the READ score was explained by the component scores (Supplementary Table 3). The per-component variance attributions were similar to those of total BARS, with additional attribution weight placed on the oculomotor and speech scores.

### F. Effect of Saccade Classes on the READ Score

To assess the importance of extracting features for each saccade class (progressive saccades, regressions, and sweeps), we computed a set of 13 features from the set of fixations and the set of all saccades. These features were computed identically to those described in the methods, with the exception of saccades being treated as a single set and the removal of the three features capturing the percentage of each saccade class. The subsequent modeling and READ score assessment methodology remained unchanged.

The READ score produced by the ablated model demonstrated degraded performance with respect to all evaluation statistics. Though still strong, the ablated READ score had a weaker correlation with BARS (r=0.77, p<0.001) and reduced reliability (ICC=0.93, p<0.001). The correlation with BARS oculomotor was slightly weaker (r=0.50, p<0.001) and the correlation with BARS speech was notably weaker (r=0.66, p<0.001). Though the ablated READ score demonstrated only a slightly diminished capacity to distinguish between controls and participants with ataxia (AUC=0.86, p<0.001), the ability to detect subclinical oculomotor signs (AUC=0.64, p=0.09) and responsiveness to disease progression (d=0.28, p=0.07) were both adversely impacted and no longer significant. The relative importance of features in the original model and this experimental removal of features specific to each saccade class convey the importance of characterizing the kinematics of sweeps in addition to the kinematics of progressive and regressive saccades.^14^

### G. Association between the READ Score and VAQ

Though some questions on VAQ ask about reading, they are designed to relate to factors such as visual acuity (e.g., trouble reading small print) or visual processing speed (e.g., trouble reading moving text) as opposed to the difficulty of reading itself. We did, however, observe a weak correlation between visual processing speed and the READ score (r=0.27, p=0.01), which may reflect the increased fixation duration, regression percentage, and number of saccades of ataxia participants during the reading task. In contrast, the visual acuity (r=0.00, p=0.99) and depth perception components (r=0.17, p=0.10) of VAQ may be more influenced by ocular motility defects^15^ that were not well-captured by the reading task. More specifically, the large font and high contrast of the passage, controlled lighting in the booth, and stationary position of the monitor may have reduced the impact of impaired visual acuity, depth perception, and vergence. Future work varying these controlled factors may lead to measures that more closely reflect these particular aspects of visual function.

## Supplementary Figures

**Supplementary Figure 1.**
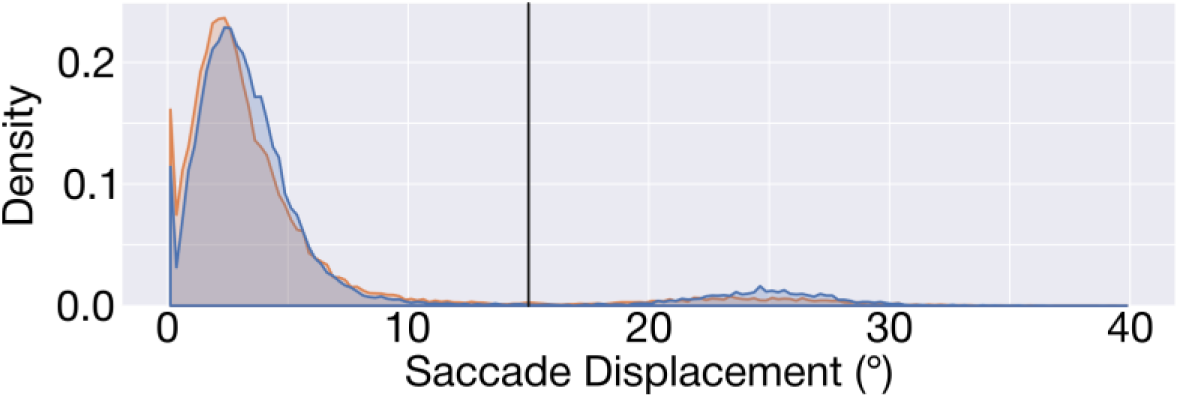
Probability density of saccade displacements. The orange curve denotes data from participants with ataxia and the blue curve denotes data from healthy controls. The black vertical line denotes the chosen 15° threshold for labeling a saccade as a sweep.

**Supplementary Figure 2.**
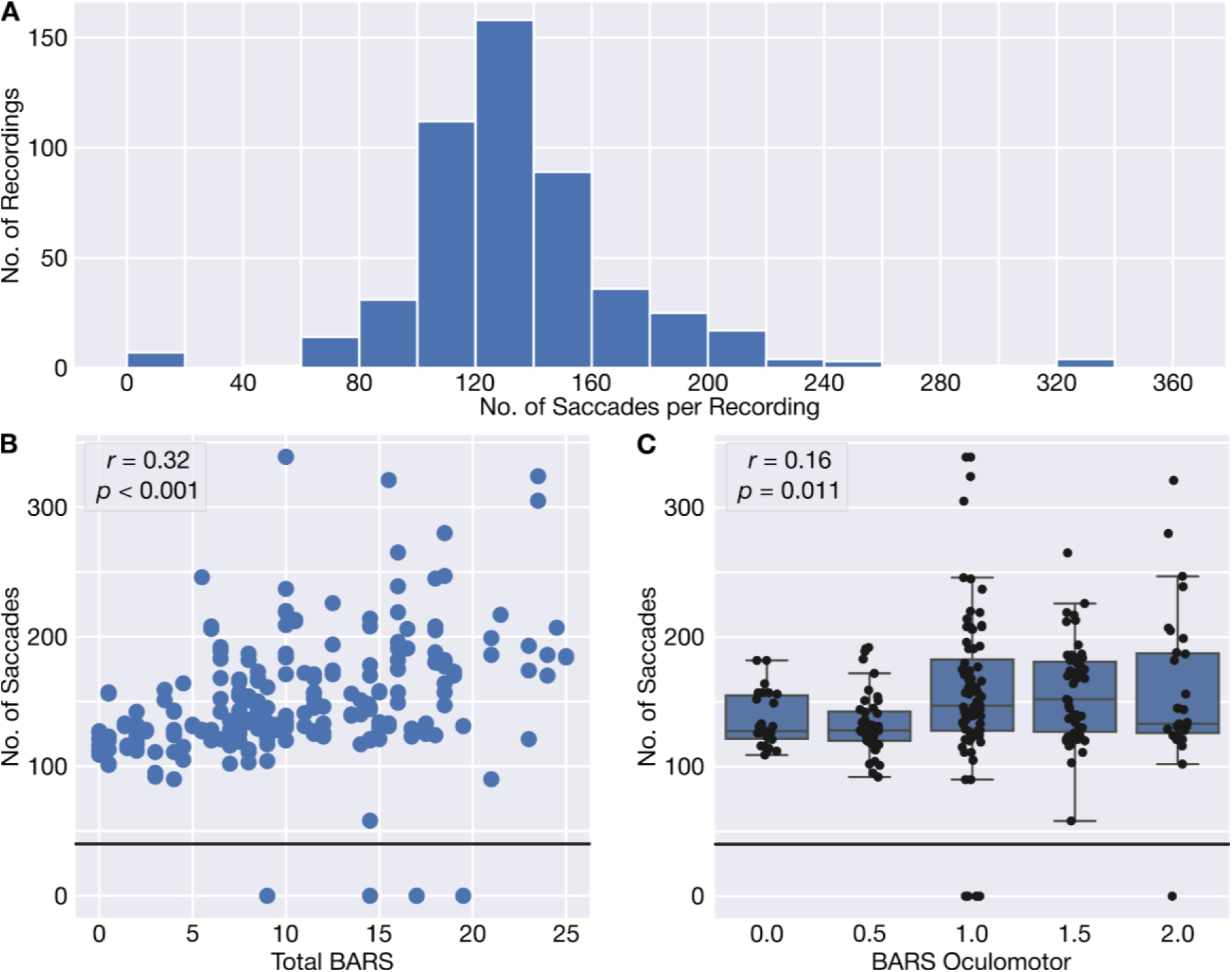
The number of saccades per task recording. (**A**) The distribution across all recordings (both participants with ataxia and healthy controls). (**B**) The relationship between clinician-assessed overall ataxia severity and the number of saccades in each task recording. The horizontal black line at 40 saccades denotes the threshold for exclusion of a recording from further analysis. (**C**) The relationship between the oculomotor component of the Brief Ataxia Rating Scale (BARS) and the number of saccades in each task recording. Box plots denote each quartile, with a maximum whisker length of 1.5 times the interquartile range. The overlaid swarm shows each task recording.

**Supplementary Figure 3.**
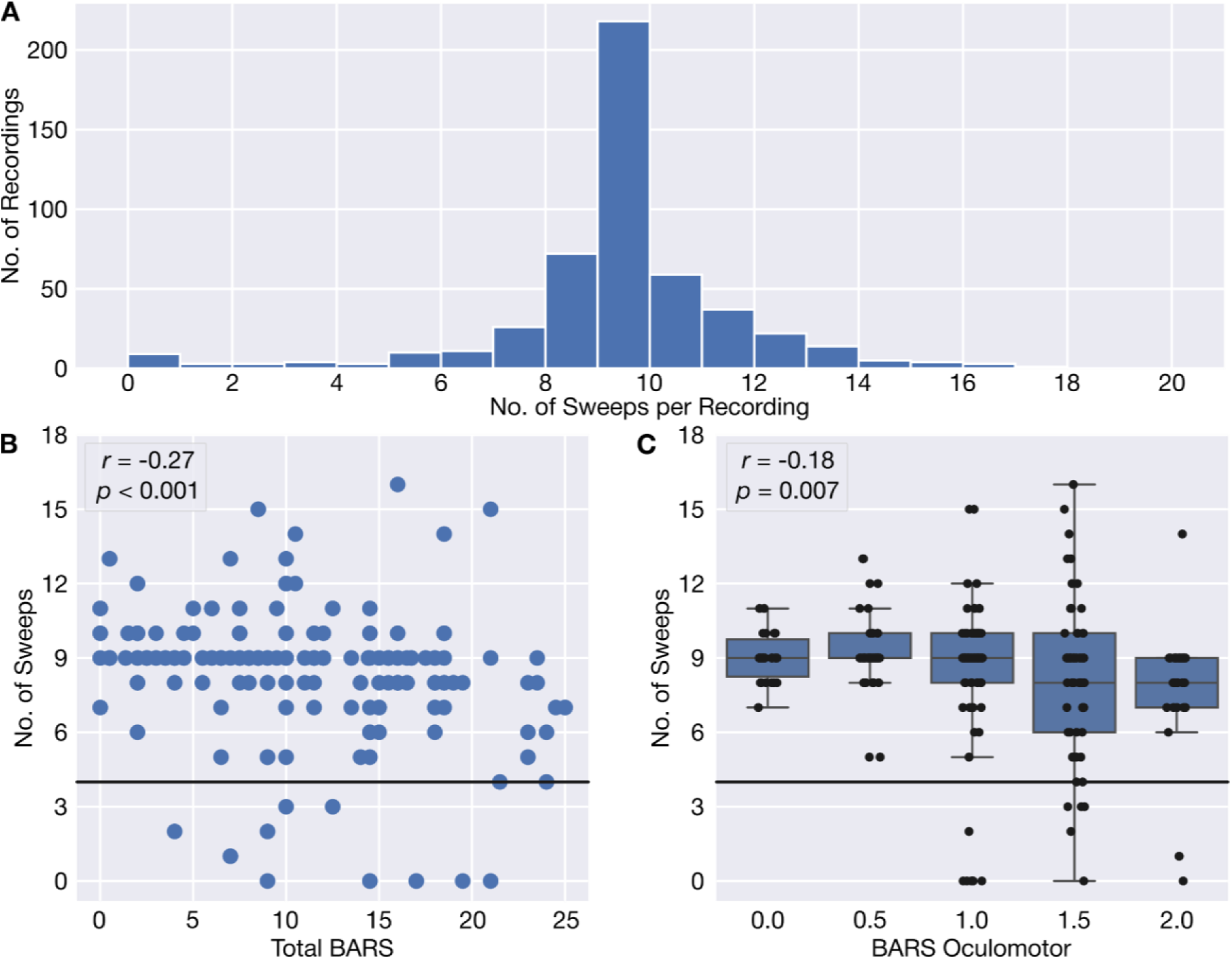
The number of sweeps (saccades with at least 15° displacement) per task recording. (**A**) The distribution across all recordings (both participants with ataxia and healthy controls). The presented passage had ten lines of text, resulting in a peak at nine sweeps. Additional sweeps may be due to regressions to previous lines or severe undersweep. Fewer sweeps result from data quality filters (e.g., exclusion due to adjacent blinks). (**B**) The relationship between clinician-assessed overall ataxia severity and the number of sweeps in each task recording. The horizontal black line at four sweeps denotes the threshold for exclusion of a recording from further analysis. (**C**) The relationship between the oculomotor component of the Brief Ataxia Rating Scale (BARS) and the number of sweeps in each task recording. Box plots denote each quartile, with a maximum whisker length of 1.5 times the interquartile range. The overlaid swarm shows each task recording.

**Supplementary Figure 4.**
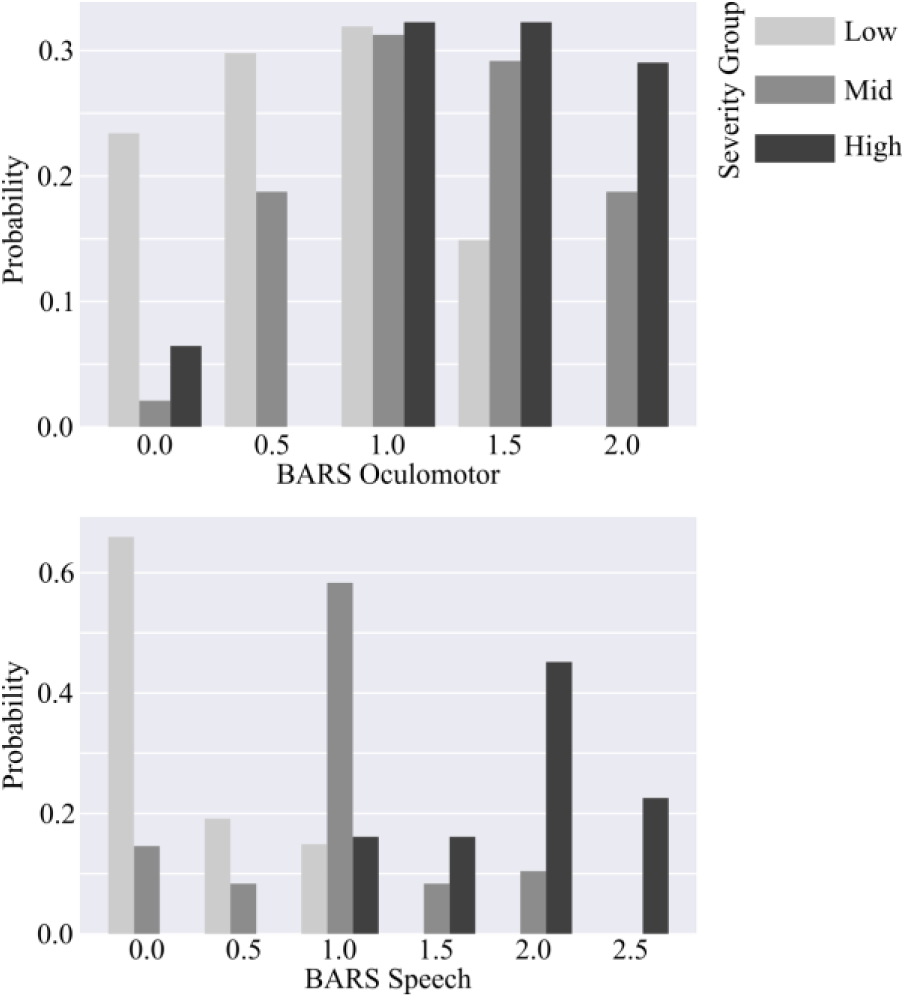
Distribution of each severity group, defined on the total Brief Ataxia Rating Scale (BARS), with respect to the speech and oculomotor components of BARS.

**Supplementary Figure 5.**
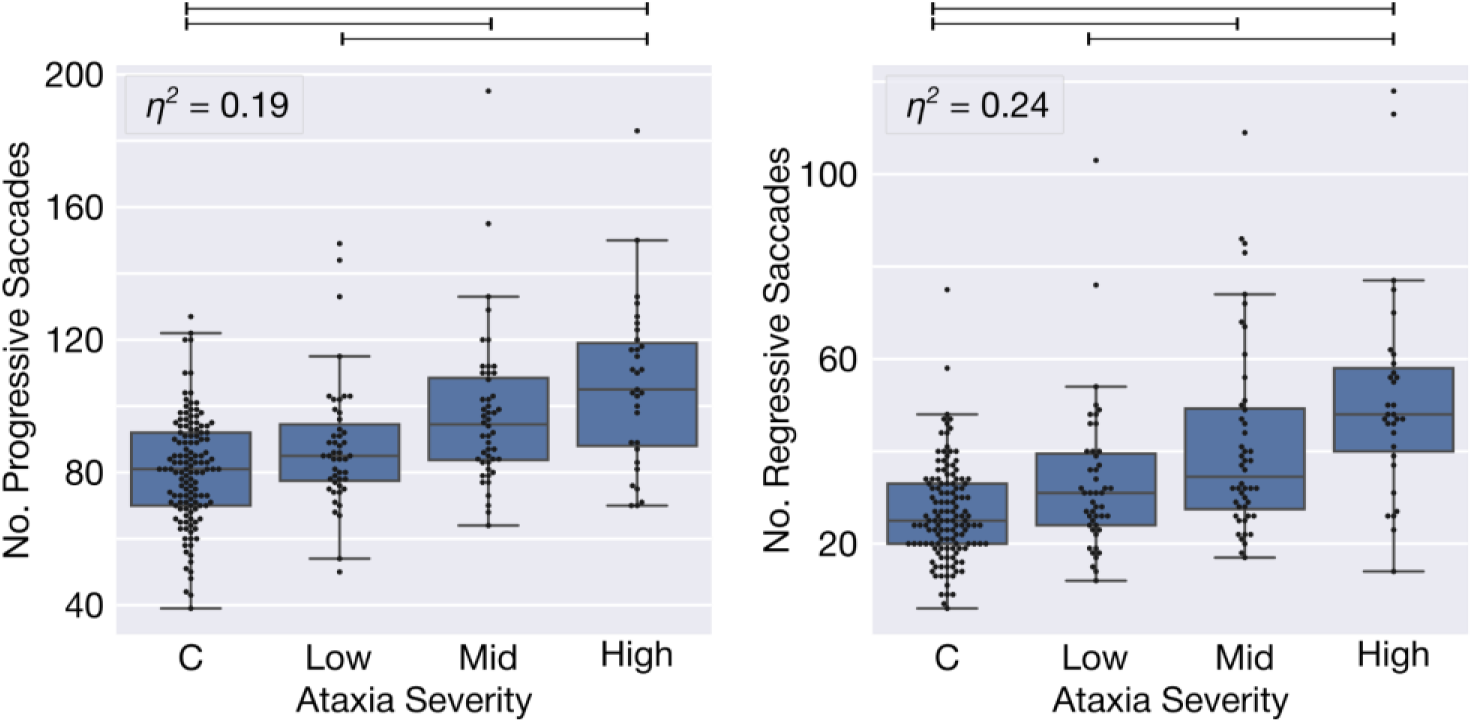
The number of progressive and regressive saccades per task recording, stratified by disease severity. These measures are calculated and presented in the same manner as the kinematic measures in Fig. 2. Both have *p*-values less than 0.001.

**Supplementary Figure 6.**
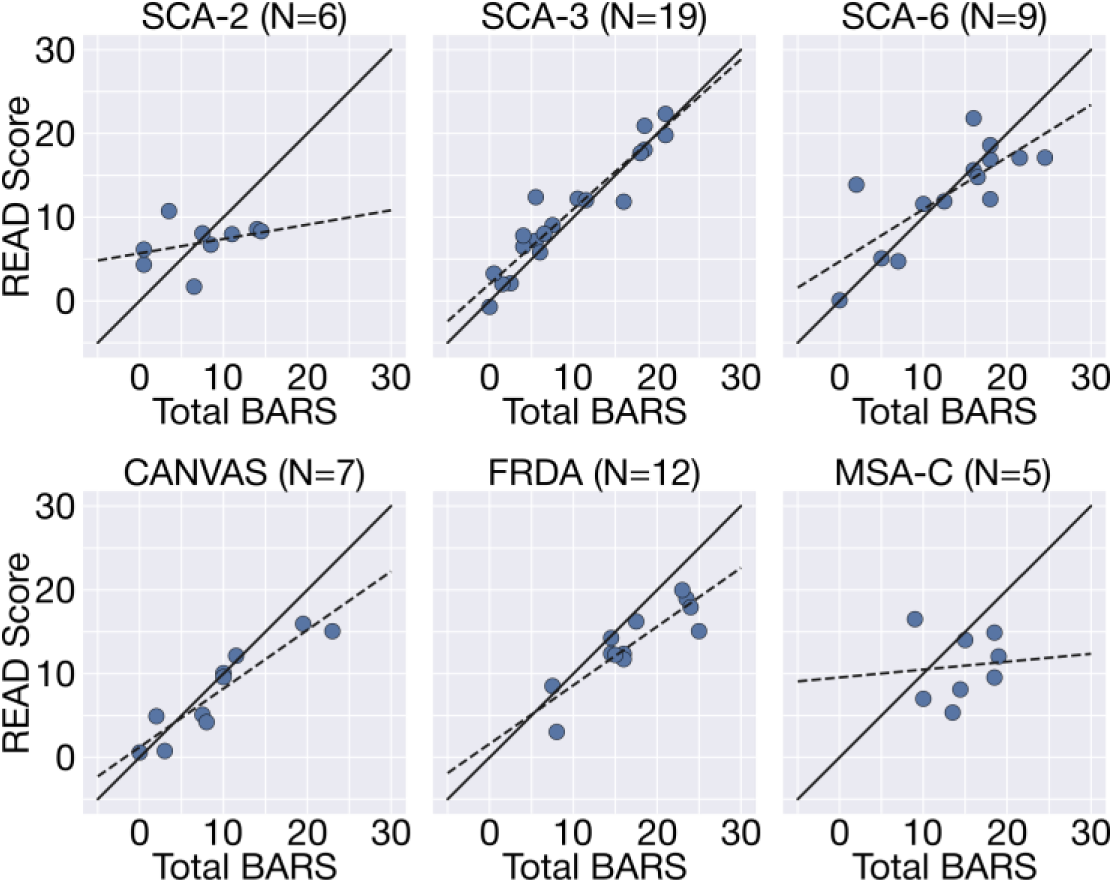
Per-recording READ score versus Brief Ataxia Rating Scale (BARS) for the six most frequent ataxia diagnoses in the data set. Each point represents a single task recording (participants could have multiple task recordings from different timepoints). The solid black line denotes perfect agreement with total Brief Ataxia Rating Scale (BARS) and the dotted black line denotes a least-squares regression. Abbreviations: SCA—Spinocerebellar ataxia; CANVAS—Cerebellar Ataxia, Neuropathy, and Vestibular Areflexia Syndrome; FRDA—Friedreich’s Ataxia; MSA-C—Multiple System Atrophy (Cerebellar Type)

**Supplementary Figure 7.**
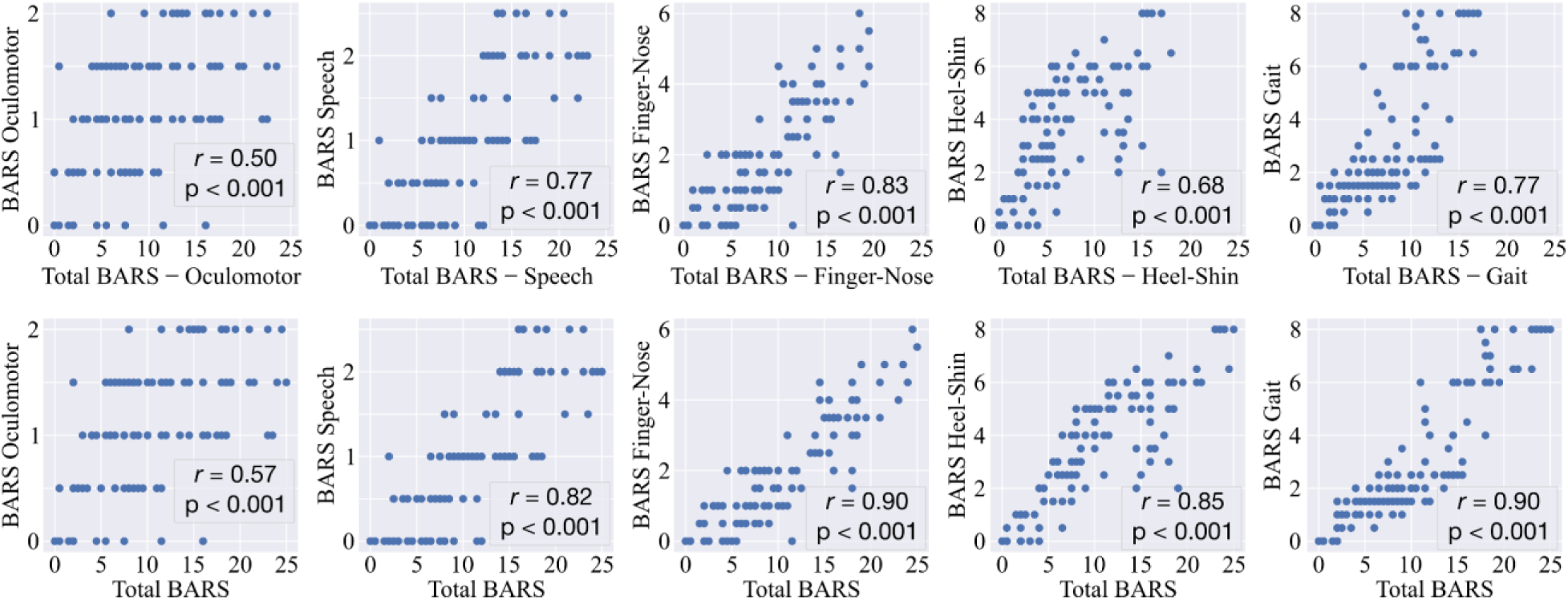
Relationship between each Brief Ataxia Rating Scale (BARS) subdomain, the remainder of BARS (top), and total BARS (bottom).

## Supplementary Tables

**Supplementary Table 1.**
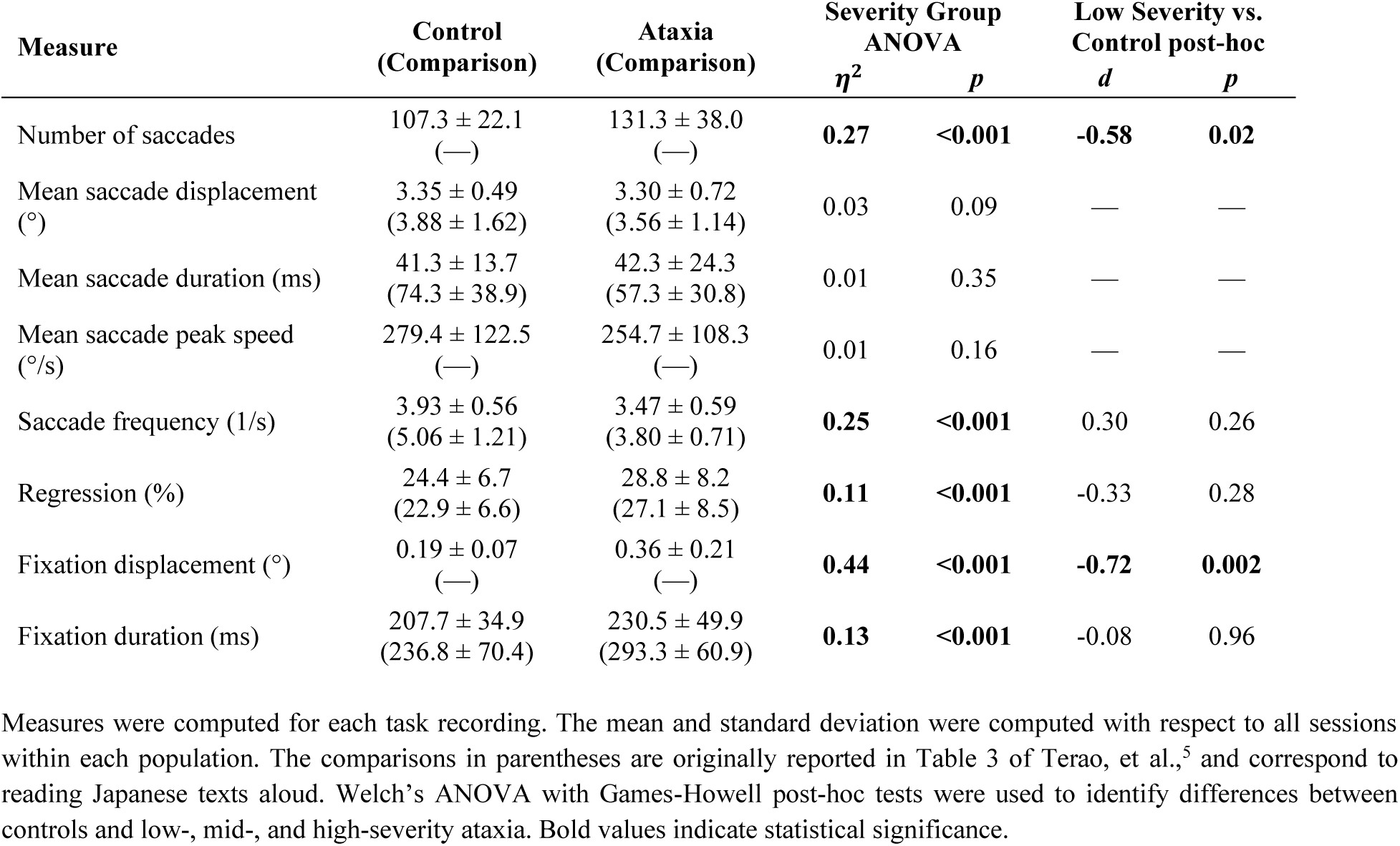
Kinematic Measures of Saccades and Fixations

**Supplementary Table 2.** READ Score Feature Importance

| Subset | Feature | Importance |
| --- | --- | --- |
| **Fixations** | **Displacement Mean** | **2.38 ± 0.04** |
|  | Displacement CV | 0.07 ± 0.03 |
|  | Duration Mean | 0.00 ± 0.01 |
|  | **Duration CV** | **1.25 ± 0.07** |
|  | Peak Speed Mean | -0.38 ± 0.03 |
|  | Peak Speed CV | -0.06 ± 0.06 |
| **Progressive Saccades** | Pct. Progressive Saccades | 0.00 ± 0.00 |
|  | Displacement Mean | -0.02 ± 0.04 |
|  | Displacement CV | -0.12 ± 0.07 |
|  | Duration Mean | -0.00 ± 0.02 |
|  | Duration CV | 0.00 ± 0.00 |
|  | Peak Speed Mean | -0.05 ± 0.03 |
|  | Peak Speed CV | 0.00 ± 0.01 |
| **Regressions** | **Pct. Regressions** | **1.09 ± 0.04** |
|  | Displacement Mean | 0.44 ± 0.05 |
|  | Displacement CV | 0.00 ± 0.01 |
|  | Duration Mean | 0.00 ± 0.01 |
|  | Duration CV | -0.08 ± 0.05 |
|  | Peak Speed Mean | -0.04 ± 0.04 |
|  | Peak Speed CV | 0.04 ± 0.04 |
| **Sweeps** | **Pct. Sweeps** | **-2.67 ± 0.03** |
|  | Displacement Mean | -0.03 ± 0.02 |
|  | Displacement CV | 0.40 ± 0.03 |
|  | Duration Mean | -0.70 ± 0.07 |
|  | Duration CV | 0.05 ± 0.04 |
|  | Peak Speed Mean | 0.00 ± 0.01 |
|  | Peak Speed CV | 0.41 ± 0.06 |
| **All Saccades** | **Saccade Frequency** | **-1.90 ± 0.03** |

Abbreviations: CV— Coefficient of Variation; Pct.— Percentage
The relative importance of each feature comprising the READ score was determined using the magnitude of the model coefficients. The mean and standard deviation of model coefficients reported are computed over all iterations of leave-one-subject-out cross-validation. The five most important features are in bold.

**Supplementary Table 3.** Amount of variance explained by each BARS component score

| Measure | Adjusted | BARS Component Score |  |  |  |  |
| --- | --- | --- | --- | --- | --- | --- |
| | $R^2$ | Oculomotor | Speech | Finger-Nose | Gait | Heel-Shin |
| Total BARS | 1.00 | 8.1% | 18.7% | 23.5% | 26.2% | 23.5% |
| READ Score | 0.67 | 11.4% | 23.0% | 21.5% | 25.4% | 18.7% |
| Number of saccades | 0.16 | 3.5% | 26.2% | 16.0% | 25.6% | 28.8% |
| Mean saccade displacement (°) | -0.02 | — | — | — | — | — |
| Mean saccade duration (ms) | 0.05 | 5.3% | 47.7% | 18.4% | 6.4% | 22.2% |
| Mean saccade peak speed (°/s) | 0.00 | — | — | — | — | — |
| Saccade frequency (1/s) | 0.24 | 3.5% | 23.1% | 23.9% | 37.7% | 11.7% |
| Regression (%) | 0.09 | 15.3% | 44.6% | 14.7% | 11.1% | 14.4% |
| Fixation displacement (°) | 0.36 | 41.0% | 11.9% | 15.6% | 18.2% | 13.3% |
| Fixation duration (ms) | 0.17 | 11.0% | 9.5% | 33.5% | 31.4% | 14.6% |

Measures were computed for each task recording. Multiple linear regression was performed with each component score of the Brief Ataxia Rating Scale as predictors and the respective measure as the dependent variable. The importance score is the percentage of the proportion of explained variance ( $R^2$ ) attributed to each predictor. Importance scores are not reported for measures where less than 5% of the variance was explained by the multiple linear regression model.

